# Prediction of MCI-to-AD progression with atrophy-weighted standard uptake value ratios of ^18^F-Florbetapir PET

**DOI:** 10.1101/2023.03.09.23287059

**Authors:** Yu-Hua Dean Fang, Jose U. Perucho, Sheng-Chieh Chiu, Yun-Chi Lin, Jonathan E. McConathy

**Affiliations:** Department of Radiology, University of Alabama at Birmingham; Department of Biomedical Engineering, University of Alabama at Birmingham

## Abstract

Accurate prediction of MCI-to-AD progression is an important yet challenging task. We introduce a new quantitative parameter: the atrophy-weighted standard uptake value ratio (awSUVR), defined as the PET SUVR divided by the hippocampal volume measured with MR, and evaluate whether it may provide better prediction of the MCI-to-AD progression. Materials and Methods: We used ADNI data to evaluate the prediction performances of the awSUVR against SUVR. 571, 363 and 252 18-F-Florbetaipir scans were selected based on criteria of conversion at the third, fifth and seventh year after the PET scans, respectively. Corresponding MR scans were segmented with Freesurfer and applied on PET for SUVR and awSUVR computation. We also searched for the optimal combination of target and reference regions. In addition to evaluating the overall prediction performances, we also evaluated the prediction for APOE4 carriers and non-carriers. For the scans with false predictions, we used 18-F-Flortaucipir scans to investigate the potential source of error. Results: awSUVR provides more accurate prediction than the SUVR in all three progression criteria. The 5-year prediction accuracy/sensitivity/specificity is 90/81/93% for awSUVR and 86/81/88% for SUV. awSUVR also yields good 3- and 7-year prediction accuracy/sensitivity/specificity of 91/57/96 and 92/89/93, respectively. APOE4 carriers generally are slightly more difficult to predict for the progression. False negative prediction is found to either due to a near-cutoff mis-classification or potentially non-AD dementia pathology. False positive prediction is mainly due to the slightly delayed progression than the expected progression time. Conclusion: We demonstrated with ADNI data that 18-F-Florbetapir SUVR weighted with hippocampus volume may provide good prediction power with over 90% accuracy in MCI-to-AD progression.

## Introduction

Alzheimer’s disease (AD) is the most common form of neurodegenerative dementia. With recent advancement in AD drugs and the FDA approval of Amyloid-targeting drugs, treating AD patients has become a reality and redefined many aspects of the management of AD. In current schemes of Amyloid-targeting drugs, it is thought that the treatment shall happen before the actual onset of AD, preferably at the mild cognitive impairment (MCI) stage. However, 50-80% of all MCI patients will convert to AD dementia^1, 2^ so not every MCI subject shall undergo Amyloid-clearing therapies. Moreover, current Amyloid-targeting drugs have been found to be associated with amyloid-related imaging abnormalities (ARIA) that typically present as brain edema or hemorrhage. Although such side effects are usually temporary and not life-threatening, they do present a risk to the patients who are often older adults that could be more vulnerable to ARIA. Accordingly, correct identification of MCI patients that possess a high risk of progression to AD may play a crucial role in AD therapies by maximizing the treatment efficacy and reducing risks posed by the unnecessary treatment given to stable MCI patients.

Various attempts have been done to develop prediction model to evaluate the likelihood of an MCI subject to progress into AD at given time windows. Imaging-based models, either univariate or multivariate, have been a popular approach for such prediction tasks. Depending on the criteria of MCI-to-AD criteria and timepoint, the current state of art models generally yields approximately 80% accuracy^3-9^. A higher prediction accuracy will be clinically desirable for decision making, both for the clinicians and patients. In the case of AD, a balanced prediction sensitivity and specificity is also a desirable feature to achieve the maximal balance between risk and benefits. Amyloid PET scans have been shown to be sensitive but less specific to predict for the MCI-to-AD progression. Therefore, improvement towards a high accuracy while maintaining a balanced sensitivity and specificity is a still challenging yet critical topic of research.

The other important aspect that has been less examined is the effect of APOE4 carriage on the prediction model. It is well known that APOE4 carriers present a higher risk of developing AD as well as progressing from MCI to AD, so there might be a high demand for therapeutics to be applied over APOE4+ MCI patients. On the other hand, current studies also show that APOE4 carriers are much more likely to develop ARIA when they receive the Amyloid-targeting drugs compared to non-carriers^10^. Accordingly, a proper examination of the prediction power over APOE4 carriers is of great importance so the physicians and patients may better evaluate the risk and benefits before initiating the Amyloid-targeting therapies.

We aim to develop a univariate prediction model based on PET and MRI scans for accurate prediction of MCI-to-AD progression. A new parameter, atrophy-weighted SUVR (awSUVR) of ^18^F-FLorbetapir is proposed and tested for better prediction power over the conventional SUVR. Retrospective data from ADNI were used to test the proposed parameter, determine the best image processing approach, and evaluate the prediction performances for MCI-to-AD progression. Moreover, we also examined the ^18^F-Flortaucipir scans of the subjects that were falsely predicted through our model to understand the potential reasons of misprediction.

## Materials and methods

### Subject selection

Data used in the preparation of this article were obtained from the ADNI database (adni.loni.usc.edu). The ADNI was launched in 2003 as a public-private partnership, led by Principal Investigator Michael W Weiner, MD. Data collection and sharing for this project was funded by the Alzheimer’s Disease Neuroimaging Initiative (ADNI) (National Institutes of Health Grant U01 AG024904) and DOD ADNI (Department of Defense award number W81XWH-12-2-0012). ADNI is funded by the National Institute on Aging, the National Institute of Biomedical Imaging and Bioengineering, and through contribution of multiple entities.

Clinical diagnosis and cognitive function evaluation were performed according to the ADNI procedure manuals. For each subject, diagnoses of cognitively normal, MCI or AD were made during the baseline evaluation and throughout the follow-ups. We first identified eligible ^18^F-Florbetapir scans taken at the MCI stage with the following criteria: (1) There must be an MCI diagnosis within 90 days of the ^18^F-Florbetapir scan OR the nearest diagnoses made before and after the ^18^F-Florbetapir scan are both MCI, (2) There must be an MRI study within one year of the ^18^F-Florbetapir scan, and (3) There must be a preprocessed PET dataset named ‘AV45_Coreg_Avg_Std_Img_and_Vox_Siz_Uniform_6mm_Res’ that was available for downloading from ADNI website. Next, we defined a ‘converter’ scan under a specific x-year criteria by: (1) Within x years after the ^18^F-Florbetapir scan, the subject’s diagnosis had converted from MCI to AD, (2) after the MCI-to-AD conversion, there was at least another diagnosis made during followups, and (3) after the MCI-to-AD conversion, all diagnoses remained as AD. Those who reverted from AD back to MCI were excluded in this study. For the ‘non-converter’ scans under a specific x-year criteria, it was defined as (1) After x years past the ^18^F-Florbetapir scan, there must be at least another MCI diagnosis made for the subject, and (2) Between this MCI diagnosis and the ^18^F-Florbetapir scan, all diagnoses made in between must remain as MCI.

We wish to note that we are referring to converters and non-converters based on scans, not individual subjects, in order to maintain a sufficiently large sample size. When a subject has multiple scans that are eligible for either a converter’s or a non-converter’s criteria, each scan is treated as an independent entity. All data were treated as cross-sectional measurements. It is therefore possible that a single subject possesses both converter and non-converter scans.

### Image acquisition and processing

PET and MRI were acquired based on the ADNI protocols. We used the standard T1-weighted images for the MR-based image processing and the static images summed over 50 to 70 minutes post inject for the PET data. All PET images were post-processed and filtered by scanner-specific filter functions to reach the uniform and isotropic spatial resolution of 6mm FWHM^11^. We used the ‘AV45_Coreg_Avg_Std_Img_and_Vox_Siz_Uniform_6mm_Res’ PET images downloadable for each subject from the ADNI website. This PET image was first corrected for the partial volume effect with the Reblurred Van-Cittert method under the PETPVC toolbox^12^ by assuming a 6mm FWHM. The structural T1-weighted images based on MP-RAGE or SPGR sequences were processed with FreeSurfer v7.3^13^ for brain parcellation. The Tl-weighted images for a specific PET scan was co-registered to PET with the ‘mri_coreg’ function in FreeSurfer v7.3. The estimated rigid-body transformation was then applied on the FreeSurfer-segmented brain regions to align those delineated regions with the PET scan. SUVR was then calculated with the partial volume-corrected images for the segmented volumes of interest (VOIs) under a given set of target and reference regions.

### Atrophy-weighted SUVR

We define the new PET parameter awSUVR as the PET SUVR divided by the hippocampal volume. The hippocampal volume was calculated through the Freesurfer parcellation followed by a dedicated hippocampal subregion segmentation procedure^14^ within Freesurfer. Total hippocampal volume was calculated from the estimated probability maps of each voxel’s likelihood of being within the hippocampus. The total intracranial volume (ICV) was estimated by Freesufer after the brain parcellation^15^ and used to normalize the hippocampal volume. The awSUVR was then calculated as:

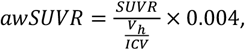

where *V*_*h*_ is the hippocampal volume estimated by Freesufer and the scaling factor of 0.004 represents the typical hippocampal:ICV volume ratio.

### Selection of target and reference region VOIs

We used a set of VOI from literature to serve as a ‘standard’ VOI set and a search method to determine the optimal VOI choices under the given discriminative task. The target VOI in the standard VOI set was a cortical composite VOI including the frontal, anterior/posterior cingulate, lateral parietal, lateral temporal regions^16, 17^ and the reference region VOI was the whole cerebellum. The search method, on the other hand, tries to find the best combination of target VOIs and reference region VOIs that yields the best prediction performances of MCI-to-AD progression. When searching for the optimal VOIs for an x-year conversion criteria, an ROC analysis was repeatedly performed with combinations of three subregions from the frontal, temporal, cinulate and lateral parietal regions as the target region. The reference region would be either the whole cerebellum, eroded white matter^18^, or the combination of the cerebellum and the eroded white matter. All possible combinations of the three subregions and the SUVR and awSUVR calculation and used in the ROC analysis which would calculate the accuracy, sensitivity and specificity for all the VOI combinations. After that, we then tested all possible combinations of four subregions and all possible combinations of five subregions under the three choices of reference regions. In the end, the optimal VOI set was defined as the combination of target and reference region choices with the highest accuracy. If more than one combination yielded the highest accuracies, the combination with the highest sensitivity was selected as the optimal VOI set. This VOI selection procedure is denoted as ‘VOI optimization’ in this work.

### ROC analysis

In this study, we tested three different conversion criteria: 3-yr, 5-yr and 7-yr progression after the PET scan. For each criteria, we perform ROC analysis with the following parameters: hippocampal volume normalized by ICV, SUVR and awSUVR. For SUVR and awSUVR, we tested the ROC performances with the standard VOI set and with the VOI-optimization procedure. In addition, we also tested the prediction performances when a SUVR cutoff of 1.11 ^19, 20^ was used and calculated the accuracy/sensitivity/specificity.

### Examination of falsely predicted cases

We took the 5-yr progression prediction model that yielded the best prediction accuracy and identified the subjects that were falsely predicted. For each of these subjects, we examined the following characteristics. First, for false positive cases, we examined the duration of follow-ups and whether there were records of MCI-to-AD progression after five years. We also checked if there were ^18^F-Flortaucipir scans available after five years past the ^18^F-Florbetapir scans. The tau SUVR was retrieved from the ^18^F-Flortaucipir analysis performed by UC Berkeley and Lawrence Berkeley National Laboratory that was downloaded from ADNI as ‘UC Berkeley - AV1451 8mm Res Analysis’ with version of 2023-02-17. The meta-temporal VOI was used as the target region with the inferior cerebellar reference region^21^. The cutoff for ^18^F-FLortaucipir was 1.23 to determine the tau positivity^21^. Second, for the false negative cases, we examined whether it is due to the ‘gray zone’ near cutoff values as previously shown in cases of ^18^F-Florbetapir. We also examined whether the ^a^-Flortaucipir showed positivity under the UC Berkeley analysis in those cases.

## Results

After screening for the eligible scans based on the selection criteria, we have identified 571, 363 and 252 eligible scans for the 3-, 5- and 7-yr conversion criteria, respectively. Among those scans, 82 (14%), 102 (28%) and 114 (45%) are converters under the 3-, 5- and 7yr conversion criteria, respectively. Demographics of the subjects was summarized in Table 1. The prediction performances of hippocampal volume, SUVR and awSUVR were summarized in Table 2. In all the three conversion criteria, awSUVR has outperformed SUVR by yielding a higher accuracy. For example, when using SUVR under VOI optimization for predicting a five-year conversion, the optimal prediction performance is 86/81/88% in accuracy/sensitivity/specificity. awSUVR improved the results by increasing the accuracy to 90% and the specificity to 93%. Moreover, awSUVR has achieved 90% or more prediction accuracy in all three time points of progression. In general, a better balance between the sensitivity and specificity was also observed with awSUVR results. The other finding is that the prediction has been more accurate in APOE4 non-carriers although the difference between APOE4+ and APOE4-cohorts was not dramatically different. awSUVR has been able to achieve a better balance between sensitivity and specificity, especially in the APOE4-group. Table 3 summarized the optimal VOI identified through the VOI optimization procedure for SUVR and awSUVR under the different criteria.

**Table 1.** Demographics of the subjects and their scans used for the three prediction criteria

| Clinical Characteristics | 3-year conversion cohort | 5-year conversion cohort | 7-year conversion cohort |
| --- | --- | --- | --- |
| Total n (scans) | 571 | 363 | 252 |
| Gender |  |  |  |
| *Male* | 322 | 205 | 143 |
| *Female* | 249 | 158 | 109 |
| Age | 73.916 ± 7.685 | 73.608 ± 7.594 | 73.545 ± 7.636 |
| Education | 16.322 ± 2.700 | 16.413 ± 2.681 | 16.520 ± 2.583 |
| MMSE | 26.747 ± 3.325 | 26.273 ± 3.746 | 25.627 ± 4.037 |
| APOE<sup>†</sup> |  |  |  |
| 3 | 350 | 218 | 139 |
| 4 | 170 | 110 | 82 |
| 5 | 51 | 35 | 31 |

<sup>†</sup> APOE status of 3 denotes that both genes have the *e2* or *e3* alleles, 4 denotes that one gene has the *e4* allele, and 5 denotes that both genes have the *e4* allele.

**Table 2.** Prediction performances of MR-based and PET-based features.

| Task | Parameter | Target VOI | Reference VOI | Cutoff value | All subjects |  | APOE4+ | APOE- |
| --- | --- | --- | --- | --- | --- | --- | --- | --- |
|  |  |  |  |  | AUC | Acc/Sen/Spe | Acc/Sen/Spe | Acc/Sen/Spe |
| 3-yr progression prediction | Hippocampal volume |  |  | 0.0027 | 0.75 | 86/2/100 | 75/0/100 | 93/4/100 |
|  | SUVr (1.1 cutoff) | Cortical composite | Cerebellum | 1.1 |  | 68/91/64 | 54/98/39 | 77/78/77 |
|  | SUVr | Cortical composite | Cerebellum | 1.659 | 0.86 | 88/35/97 | 80/36/94 | 93/33/98 |
|  | SUVr | Optimal search | Optimal search | 0.842 | 0.86 | 90/43/98 | 84/47/96 | 94/33/99 |
|  | awSUVr | Cortical composite | Cerebellum | 1.638 | 0.89 | 89/56/95 | 82/55/91 | 94/59/97 |
|  | **awSUVr** | **Optimal search** | **Optimal search** | **1.165** | **0.90** | **91/57/96** | **85/60/93** | **94/52/98** |
| 5-yr progression prediction | Hippocampal volume |  |  | 0.0038 | 0.80 | 67/85/60 | 76/85/72 | 54/96/38 |
|  | SUVr (1.1 cutoff) | Cortical composite | Cerebellum | 1.1 |  | 76/90/70 | 71/98/49 | 79/76/80 |
|  | SUVr | Cortical composite | Cerebellum | 1.253 | 0.88 | 84/82/84 | 80/91/71 | 86/68/90 |
|  | SUVr | Optimal search | Optimal search | 0.649 | 0.88 | 86/81/88 | 87/94/81 | 86/59/91 |
|  | awSUVr | Cortical composite | Cerebellum | 1.435 | 0.92 | 89/74/95 | 84/75/91 | 92/70/96 |
|  | **awSUVr** | **Optimal search** | **Optimal search** | **0.888** | **0.93** | **90/81/93** | **89/86/91** | **91/73/94** |
| 7-yr progression prediction | Hippocampal volume |  |  | 0.0039 | 0.82 | 74/85/66 | 76/69/82 | 70/91/53 |
|  | SUVr (1.1 cutoff) | Cortical composite | Cerebellum | 1.1 |  | 85/89/82 | 85/99/62 | 85/72/91 |
|  | SUVr | Cortical composite | Cerebellum | 1.199 | 0.92 | 88/85/91 | 88/94/79 | 88/70/97 |
|  | SUVr | Optimal search | Optimal search | 1.223 | 0.92 | 91/88/93 | 92/97/83 | 90/72/98 |
|  | awSUVr | Cortical composite | Cerebellum | 1.126 | 0.95 | 89/90/88 | 87/92/79 | 91/88/93 |
|  | **awSUVr** | **Optimal search** | **Optimal search** | **0.785** | **0.95** | **92/89/93** | **88/90/83** | **95/88/98** |

**Table 3.** The optimal VOI combination identified for prediction tasks

| Task | Feature | Reference region | Target region (Freesurfer label) |
| --- | --- | --- | --- |
| 3-yr progression prediction | SUVR | Eroded WM | medialorbitofrontal, rostralmiddlefrontal, entorhinal |
| 3-yr progression prediction | awSUVR | Eroded WM plus whole cerebellum | parsorbitalis, rostralmiddlefrontal, temporalpole, precuneus |
| 5-yr progression prediction | SUVR | Eroded WM | medialorbitofrontal, parstriangularis, fusiform, temporalpole, precuneus |
| 5-yr progression prediction | awSUVR | Eroded WM plus whole cerebellum | parsopercularis, bankssts, temporalpole, transversetemporal, isthmuscingulate |
| 7-yr progression prediction | SUVR | Whole cerebellum | parsorbitalis, bankssts, posteriorcingulate, supramarginal |
| 7-yr progression prediction | awSUVR | Eroded WM plus whole cerebellum | superiorfrontal, inferiortemporal, caudalanteriorcingulate |

Table 4 summarized the scans with false positive prediction with awSUVR under a five-year criteria. Out of the 17 false-positive scans, 2 (12%) converted to AD between 5^th^ and 6^th^ year, 4 (24%) converted between 6-7 years and 2 (12%) converted between 7-8 years. For those without any AD conversion recorded, two subjects had ^18^F-Flortaucipirt data after the 5^th^ year past the Amyloid PET scan and both of them showed tau positivity between the 5-6 year intervals. In total, ten out of the 17 false positive cases either converted to AD or show tau positivity between the 5 to 8 years’ period after the PET scan.

**Table 4.** Summary of the cases with a false positive prediction from awSUVR under the 5-year criteria

| Subject # | PET date | Follow-up duration after PET | Conversion date | Conversion time after PET (yr) | Tau positivity (if tau PET is available) |
| --- | --- | --- | --- | --- | --- |
| 1 | Sep, 2011 | 6.3 | Dec, 2017 | 6.3 |  |
| 2 | Sep, 2010 | 6.7 |  |  | Pos (Oct, 2015)<br>Pos (Sep, 2016) |
| 3 | Oct, 2012 | 5.9 |  |  |  |
| 4 | Oct, 2010 | 5.0 |  |  |  |
| 5 | Dec, 2010 | 6.8 |  |  |  |
| 6 | Aug, 2011 | 9.4 | Jan, 2021 | 9.4 |  |
| 7 | Aug, 2013 | 7.4 | Jan, 2021 | 7.4 |  |
| 8 | Jul, 2011 | 5.1 |  |  |  |
| 9 | Aug, 2015 | 7.0 |  |  |  |
| 10 | Aug, 2011 | 7.1 | Sep, 2017 | 6.1 |  |
| 11 | Aug, 2011 | 7.4 | Oct, 2017 | 6.1 | Pos (Dec, 2018) |
| 12 | Oct, 2011 | 5.0 |  |  |  |
| 13 | Dec, 2011 | 8.2 | Nov, 2017 | 5.9 |  |
| 14 | Mar, 2012 | 6.9 |  |  | Pos (Mar, 2018) |
| 15 | Jul, 2014 | 7.2 | Oct, 2021 | 7.2 |  |
| 16 | Sep, 2013 | 7.7 | Oct, 2019 | 6.1 |  |
| 17 | Nov, 2012 | 5.4 | Apr, 2018 | 5.4 |  |

Table 5 summarized the 19 scans with false negative prediction with awSUVR under the five-year criteria. 5 (26%) of these cases possess an awSUVR above or equal to 90% of the cutoff value of 0.89. 9 (47%) of these cases possess an awSUVR above or equal to 85% of the cutoff value. In these false negative cases, 10 (53%) have had ^18^F-Flortaucipir data after the conversion and 6 (32%) of those ^18^F-Flortaucipir scans showed tau positivity while the rest were tau negative. awSUVR was highly associated with the tau positivity.

**Table 5.** Summary of the cases with a false negative prediction from awSUVR under the 5-year criteria

| Subject # | PET date | Conversion date | awSUVR | awSUVR<br>ratio to<br>cutoff (%) | Tau positivity |
| --- | --- | --- | --- | --- | --- |
| 1 | Feb, 2013 | Jan, 2014 | 0.77 | 87 |  |
| 2 | Mar, 2013 | Mar, 2014 | 0.78 | 88 |  |
| 3 | Apr, 2013 | Apr, 2014 | 0.68 | 76 |  |
| 4 | Feb, 2014 | Oct, 2017 | 0.87 | 98 | Pos (Nov, 2017) Pos (Feb, 2019) Pos (Feb, 2020) |
| 5 | Nov, 2012 | Oct, 2014 | 0.73 | 82 |  |
| 6 | Jul, 2011 | Jul, 2012 | 0.76 | 86 |  |
| 7 | Feb, 2016 | Jan, 2019 | 0.54 | 61 | Neg (Feb, 2020) Neg (Jan, 2022) |
| 8 | Jan, 2018 | Jan, 2019 | 0.51 | 58 | Neg (Feb, 2020) Neg (Jan, 2022) |
| 9 | Sep, 2013 | Jul, 2017 | 0.81 | 91 |  |
| 10 | Sep, 2015 | Mar, 2018 | 0.63 | 71 | Neg (Jun, 2018) |
| 11 | Sep, 2017 | Aug, 2021 | 0.69 | 77 | Neg (Oct, 2021) |
| 12 | Dec, 2011 | Dec, 2013 | 0.84 | 94 |  |
| 13 | Apr, 2012 | Nov, 2012 | 0.71 | 80 | Pos (Aug, 2016) |
| 14 | May, 2012 | May, 2015 | 0.69 | 77 |  |
| 15 | Jun, 2012 | May, 2013 | 0.61 | 69 |  |
| 16 | Jun, 2012 | Jun, 2014 | 0.84 | 94 | Pos (Oct, 2017) |
| 17 | Sep, 2012 | Oct, 2016 | 0.75 | 85 | Pos (Nov, 2017) |
| 18 | Sep, 2012 | Sep, 2014 | 0.80 | 90 | Pos (Sep, 2018) Pos (Jan, 2020) |
| 19 | Jan, 2019 | Jan, 2021 | 0.65 | 73 | Pos (Jan, 2021) |

## Discussion

Development of prediction models for AD risk assessment has been an active research topic. Under current pharmaceutical advancements and FDA-approved Amyloid-targeting drugs, an accurate and reliable prediction model may facilitate a robust stratification of patients based on their individual risks of AD progression to achieve more precise and personalized treatment plans for patients with mild and early cognitive decline. A univariate cutoff-based model is easier to interpret and less prone to overfiffing compared to multivariate classifiers. In this work, we aim to evaluate the performance of ^18^F-Florbetapir in predicting the MCI-to-AD progression. In addition to the conventional SUVR, we also introduced a new PET feature awSUVR. Our data have led to several findings. First, we showed that awSUVR indeed outperformed SUVR in the progression prediction. Although such improvement was not dramatic, we have demonstrated that awSUVR has been a better predictor than SUVR in 3, 5 and 7 years for predicting the progression. This is perhaps due to the fact that, in the ‘gray zone’ subjects with near-threshold SUVR, measuring the hippocampal volume would help differentiate those with early signs of atrophy and a higher risk of progressing to AD. It is important to note that awSUVR was able to achieve 90% accuracy in all three criteria. To our best knowledge, this is by far the best prediction performance for a univariate cutoff-based model with large cohort sizes (n>250).

Second, we found that SUVR has also provided good prediction performances with an accuracy of 86% for 5-yr and above 90% for 3- and 7-yr prediction. This prediction performance of SUVR is superior than what was reported in literature. It can be due to two reasons: (1) We have applied more strict selection criteria to identify eligible scans. For example, those who reverted from AD back to MCI were excluded because there could be uncertainty for an AD diagnosis. We have also enforced the non-converters to be those without any change of diagnosis within the criteria’s period, so those with temporary progression or regression were also excluded. Such strict selection criteria may have helped us identify subjects with more accurate diagnosis which in turns improves the prediction performances. (2) We have performed VOI optimization for SUVR to identify to optimal combination of target and reference regions under a specific discriminative task. Compared to a standard VOI set, VOI optimization may help improving the prediction performance as different brain region’s Amyloid load may be of different relevance for different tasks. Our results showed that, by allowing VOI optimization, the accuracy can be improved by 2-3% for SUVR compared to the standard VOI set.e

Third, we demonstrated that awSUVR provides good prediction results for APOE4+ and APOE4-groups. Although the prediction performance in APOE4 carriers is slightly worse than the non-carriers, there is only minor differences in the prediction performances. Also, perhaps due to the data imbalance between the converters and non-converters, awSUVR-based prediction tends to be more specific than being sensitive, especially in the APOE4-group. Future studies with larger cohorts may help address whether the sensitivity can be improved with more balanced datasets.

We have also attempted to determine the reasons behind the false prediction results under a five-year progression criteria. For the false positive cases, the majority of such cases either converted between 5 and 8 years or showed tau positivity in this time interval. In other words, most of the false positive cases did convert to AD just slightly later or have been developing AD pathology. We were unable to identify why these subjects converted later than expected, but since the majority of these subjects did convert to AD soon after five years, they did own a high risk of AD progression at the time of ^18^F-Florbetapir scan and a false positive prediction may still be clinically beneficial for these subjects. On the other hand, there might be two major reasons behind the false negative cases. We speculate that about half of the false negative cases were falsely classified because their awSUVR was in the ‘gray zone’ which causes ambiguity around the cutoff value. It is likely that the Amyloid load and atrophy is approaching the detection threshold but have not yet reached the critical cutoff in such patients. For the other half false negative cases, since the awSUVR was well below cutoff and the tau SUVR were mostly below the positivity criteria, we speculate that these cases may not be with AD dementia or under the AD pathology but with other types of dementia. However, further data are necessary to confirm this hypothesis.

## Conclusion

A novel PET feature awSUVR has been proposed to quantify ^18^F-Florbetapir scans and evaluated for the performances of predicting an MCI-to-AD progression at 3, 5 and 7 years. We found that it is able to achieve more than 90% accuracy in all three criteria and to provide a good balance between sensitivity and specificity in predicting the 5- and 7-yr progression. Such prediction may help stratify MCI patients for their risks of progressing to AD and determine the optimal time for pharmaceutical intervention.

## Data Availability

All data produced in the present study are available upon reasonable request to the authors

